# Fair and Accessible Parkinson’s Disease Screening using a Machine Learning-Powered Web Platform: Research Protocol and Preliminary Results

**DOI:** 10.1101/2024.07.17.24310585

**Authors:** Peter Washington

**Affiliations:** University of Hawaii at Manoa

**Keywords:** Precision Health, Deep Learning, Self-Supervised Learning, Patient Generated Health Data

## Abstract

Digital technologies offer unprecedented opportunities to screen for conditions like Parkinson’s Disease (PD) in a scalable and accessible manner. With the widespread adoption of smartphones and computers, the general public is constantly interacting with digital interfaces, leaving behind a wealth of data that can be harnessed for health screening. Keystroke dynamics, touchscreen interactions, and other digital footprints have emerged as potential indicators of PD. By analyzing patterns in keyboard typing, touchscreen gestures, and other digital indicators, it is now possible to detect subtle motor impairments associated with PD. We propose to further develop, refine, and validate a baseline predictive model for Parkinson’s disease (PD) based on keystroke and touchscreen measurements which we have developed and tested on participants in Hawaii. Through extensive experimentation, the project aims to determine the optimal combination of features that yield the highest sensitivity and specificity in distinguishing participants with and without PD while algorithmically reducing disparities in performance across race and socioeconomic status. A central challenge of this research will be ensuring fairness by mitigating biases caused by differences in laptop and desktop screen dimensions, mouse responsiveness, and other configurations. These differences are likely to vary by socioeconomic status, requiring a thorough analysis of these disparities and employment of algorithmic fairness techniques to mitigate the underlying problem. Additionally, we will conduct human-centered design sessions to understand how to create such screening tools in a manner that is sensitive to Indigenous data sovereignty. Our findings will underscore the potential of leveraging technology-measured limb movement data as a reliable and accessible method for early detection of PD. This research holds promise for screening individuals who may potentially be affected by PD earlier in an accessible and scalable manner, thus reducing socioeconomic health disparities related to early screening and diagnosis.

## Introduction

Over 8.5 million individuals worldwide are estimated to be affected by Parkinson’s disease (PD), a neurodegenerative condition that significantly impacts a person’s motor function and daily life [1-3]. Early detection plays a crucial role in effectively managing this disease, and various data sources can be utilized for its diagnosis [4]. Notably, lower limb movements involving keyboard interactions and trackpad/touchscreen activities have proven to be reliable indicators of PD [5-7] using artificial intelligence (AI). The use of a computer as a sensor to detect PD is particularly desirable since digital devices are increasingly ubiquitous throughout the United States and therefore enable a scalable and accessible solution to PD screening.

While the use of digital consumer technologies for AI-based detection of PD is a nascent field of research which has repeatedly demonstrated high predictive power, there are a multitude of gaps which must be addressed to reduce and prevent health disparities in such AI-powered PD screening tools. First, Indigenous communities have recently maintained a general distrust towards sharing their research data because there are numerous issues pertaining to the collection, storage, analysis, and dissemination of Indigenous data by state agencies and researchers [8]. Because the extent of this mistrust towards digital data collection has yet to be evaluated for Native Hawaiians with respect to digital technologies and machine learning for healthcare purposes in particular, there is a general need to understand these sentiments in the context of Native communities. Such an understanding will help to avoid a dearth of data from these populations, an issue which would lead to disparate performances by AI models trained on those data. Second, while there are often tens or even hundreds of digitally-derived inputs fed as input to these AI models, there is a need to identify the subset of model inputs which may lead to biased performance across demographic groups. Solutions to AI bias often neglect to consider that certain model inputs may lead to bias. Third, there are differences in access to consumer technologies based on socioeconomic groups, leading to systemic differences across demographic groups in the reliability and idiosyncrasies of the mouse and keyboard movements collected. There is a critical need to mitigate these disparities, and recent innovations in the field of algorithmic fairness make this possible.

The use of artificial intelligence (AI) for Parkinson’s Disease (PD) screening is a growing field of research, with several smartphone applications and websites collecting data such as finger gestures, mouse movements, and keyboard presses to build high-performing models [9] often published in high-impact journals [9-10]. However, the study of these systems is absent of biases which can be unintentionally learned by AI models which have unequal representations of data across demographic groups. This proposal will optimize AI-based PD screening tools for fairness though the entire data pipeline: (1) creating a comfortable environment for underserved communities who were historically mistreated with respect to scientific data collection to share their digital data, helping to mitigate issues with uneven data across demographic groups; (2) optimizing the inputs that the model looks at to account for fair performance across demographic groups; and (3) training the AI models to explicitly perform equally across demographic groups, an objective which has historically been left out of AI for healthcare projects.

In the United States alone, PD affects a vast swath of the senior population, with over 90,000 new diagnoses reported annually [11]. PD primarily impacts the patient’s body and lower limbs, leading to challenges such as micrographia, drastically impeding the daily lives of those with PD [2,5]. Unfortunately, there is currently no official diagnostic procedure available for PD, resulting in numerous cases where patients remain undiagnosed or are misdiagnosed, exacerbating the difficulties associated with effective treatment [12-15]. Even unofficial diagnostic tests for PD incur substantial costs, requiring specialized equipment and laboratory tests, thus rendering the diagnostic process expensive and inconvenient [16-19]. Consequently, there is a pressing need for scalable and accessible tools for PD detection and screening. Early diagnosis of PD offers several advantages, including timely interventions and appropriate medication, enabling patients to maintain a high quality of life [20]. Parkinson’s disease (PD) is characterized by its impact on limb movements, particularly evident in lower hand movements [21-23]. Traditionally, clinical settings have been utilized for PD diagnosis, involving neurologists who consider medical history, conduct physical examinations, and observe motor movements and other distinctive symptoms [24-25].

To solve these issues, we propose the following specific aims: (1) Co-design a gamified PD screening website which is sensitive to Indigenous data sovereignty issues. We hypothesize that NHPI communities and adults over 65 more broadly will be receptive to the proposed data collection paradigm if proper safeguards are set. (2) Identify the most salient combination of features for unbiased prediction of PD status. We hypothesize that a subset of features derive from mouse movement and keyboard press data will yield optimal predictive performance of our AI model compared to using a complete set of features. (3) Mitigate racial and socioeconomic biases in the PD prediction model using algorithmic fairness approaches. We hypothesize that an algorithmic fairness approach called “fairness regularization” will lead to more even performance across demographic attributes compared to using a traditional optimization approach.

## Methods

Through the use of a readily accessible web application which can be opened on consumer devices, we will analyze both keyboard and mouse data to differentiate between participants with and without PD using AI. We have already developed this web platform, which facilitates direct performance comparison between participants through the use of identical tests. Using the collected data, we have already conducted a preliminary feasibility study which demonstrates that there are noticeable differences between participants with and without PD. We have also already developed an AI model (Figure 1) which demonstrates promising yet improvable performance in the binary classification task.

**Figure 1.**
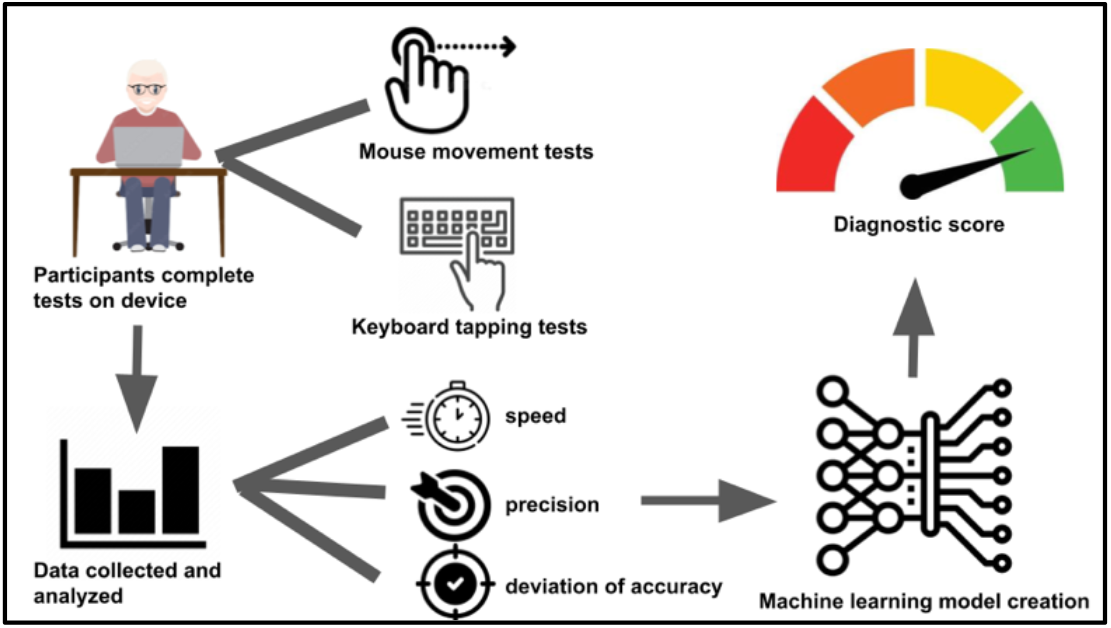
Pipeline for developing the core AI model used in this study. Mouse and keyboard data are collected from participants, a series of features are derived from the raw data, and these features are fed as input into a machine learning model with performs the prediction of PD vs. no PD.

To gather data, participants will fill out an online intake form followed by a 10-minute gamified data collection experience. Participants will be instructed to type on a keyboard while we record the corresponding timestamps and finger movements based on key positions. They will be prompted to press a specific key upon receiving a signal displayed on the screen. We will capture and store information about the expected key, the pressed key, and the time interval. This measurement process will consist of three levels, progressively increasing in difficulty by introducing greater key randomization.

For trackpad/mouse data collection, participants will be asked to hover the mouse over a designated path (Figure 2). The initial stage will involve a simple straight line to assess unintended vibrational movements. Subsequent levels will present a sine wave-like shape and later a spiral shape. Participants will be able to monitor their progress by observing a highlighted portion of the shape, while a moving light and “start”/”finish” markings will indicate the intended direction of movement. We will record and store data on the position, time, and whether the mouse is inside or outside of the shape.

**Figure 2.**
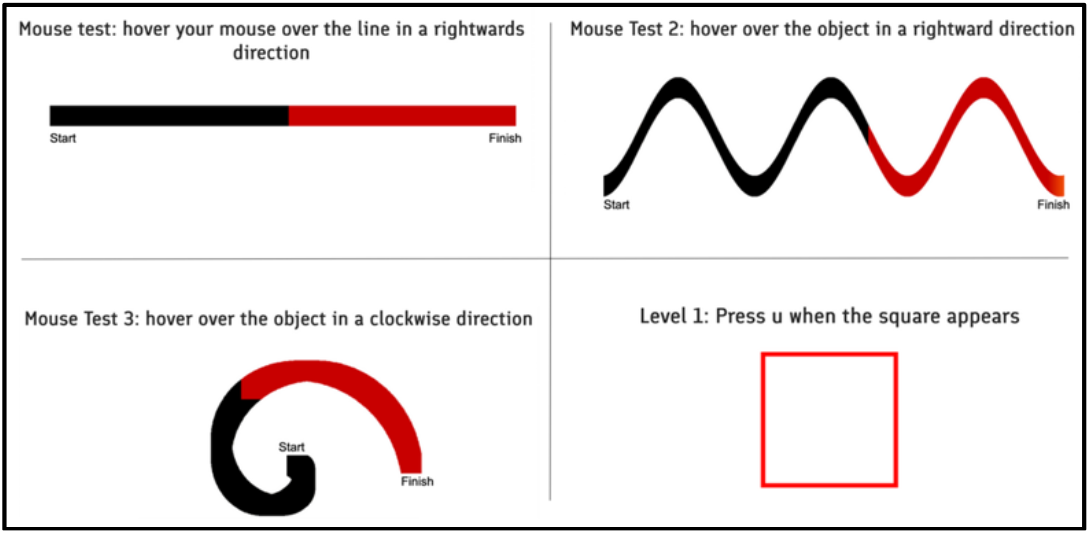
Interface of the website which houses the mouse and keyboard tests. We have already developed this website and tested it via a feasibility study. The tests consist of a combination of hovering the user’s mouse over the pre-specified shape and pressing a specified key on the keyboard when prompted.

Upon completing both the keyboard and mouse tests, the collected data will be transmitted to the database and stored using the deta.sh platform, facilitated by an API based on deta.sh micros. This will ensure secure storage and efficient management of the data. The utilization of the deta.sh platform and its associated API will guarantee robust data security and streamlined data management throughout the research process.

For Aim 1, we aim to recruit 20 participants (5 NHPI with PD, 5 NHPI without PD, 5 from another race with PD, and 5 from another race without PD). For Aims 2 and 3, we aim to recruit 200 participants (100 with and 100 without PD), matched by age and sex. For the group of 200 participants, we aim to recruit at least 20 participants from each of the following groups: Asian, White, Native Hawaiian and other Pacific Islanders, Hispanic and Latinos, Black or African American, and Native American and Alaskan Native. We similarly aim to recruit participants across the socioeconomic spectrum. We believe that the remote nature of the data collection process and the short (10-minute) nature of the study procedures makes these recruitment goals attainable. We were able to recruit 31 participants for our preliminary feasibility study with minimal effort.

For all aims, we will recruit through the Hawaii Parkinson’s Association (HPA). The Board of HPA consists of multiple individuals born and raised in Hawaii with connections to both the local PD community and the community more broadly. We will collaborate with Mr. Boster and HPA more broadly to ensure that all recruitment and study practices are inclusive and culturally appropriate. Because the entirety of the study procedures (survey and study tasks) will take less than 15 minutes to complete, retention is not an issue.

We will train 5 types of machine learning models: logistic regression, support vector machines, decision trees, random forests, and dense neural networks. For all types of models, we will perform grid search to optimize hyperparameters. For the neural networks, we will perform neural architecture search by treating the number of layers and the nodes per layer as hyperparameters. Given the dataset size and feature size, more complex neural networks are unnecessary and would likely lead to overfitting.

We aim to develop AI models that predict PD diagnostic status without bias across pre-specified attributes such as laptop and desktop screen dimensions, mouse responsiveness, and other configurations. Our hypothesis is that explicitly optimizing a model to reduce bias will outperform a model solely optimized for error reduction, the current standard in ML training. Traditionally, binary classification models minimize the error between predicted and true probability distributions using binary cross-entropy as the loss metric.

Our approach involves modifying the loss function to penalize unfair models and incorporating fairness considerations through group-based loss optimization. We will modify the binary classification setup by adding a term to the loss function which quantifies the model’s bias, leading to a loss function which is a combination of cross-entropy loss and a bias metric. We will determine a hyperparameter, C, to control the optimization’s balance between bias reduction and cross-entropy error minimization. This approach, known as “fairness optimization,” has been discussed in algorithmic fairness literature [28-31]. Specifically, we will add equalized odds and demographic parity terms to the loss function. We will incorporate fairness by partitioning the dataset into demographic groups and optimizing a weighted sum of the loss function for each group.

We will evaluate our models by comparing their performance across racial groups. We will use standard error metrics for binary classification, including accuracy, precision, recall, specificity, and ROC AUC. Additionally, we will measure bias and fairness using common metrics in the field of algorithmic fairness: equalized odds and demographic parity [28-31]. To study the impact of regularization strength (C), we will vary its value, training separate models for each setting.

## Results

This study was approved by the University of Hawaii Institutional Review Board (IRB) under protocols 2022-00857 and 2023-00948.

We have already developed the core web data collection platform shown in Figure 2. We ran a small feasibility study on 31 participants (18 without PD and 13 with PD; mean age 65.2 years +/- 10.8 years) recruited through our community partner Jerry Boster and at a presentation at the 2023 Hawaii Parkinson’s Association Symposium [32]. Five-fold cross-validation on these data yielded a mean F1-score of 77.2%. While this performance is low, we highlight the relatively tiny dataset used for this analysis. We expect that once we scale up to 200 participants, the performance will drastically increase to be aligned with prior literature in this space which regularly reports accuracy, sensitivity, and specificity above >98% when the AI models are trained on larger datasets [9].

## Discussion

While research into digital diagnostic and screening tools is becoming widely prevalent across a multitude of NIH-funded projects, including for PD disease, systematic approaches of bias mitigation strategies for such digital diagnostics is surprisingly absent despite being widely acknowledged as a pressing issue. In particular, differences in user devices are likely to vary by socioeconomic status, requiring a thorough analysis of these disparities and employment of algorithmic fairness techniques to mitigate the underlying problem.

All three of the proposed aims in this study involve an advancement of technical capabilities in the field of AI-based screening of health conditions which will lead to a reduction of bias in these systems. In Aim 1, we will advance current practices for conducting human-centered design sessions with NHPI communities. While human-computer interaction (HCI) is a mature field of study with several established techniques for co-designing with stakeholders, there is a dearth of HCI work exploring participatory design with NHPI communities for healthcare. In Aim 2, we will conduct one of the first, if not the first, study of feature selection for fairness. sAlgorithmic fairness is not traditionally studied in the context of input features. In Aim 3, we will conduct the first application of state-of-the-art algorithmic fairness algorithms to PD-based screening while tweaking these methods to our unique data modalities.

## Data Availability

All data produced in the present study are available upon reasonable request to the authors.

## Notes

### Competing Interest Statement

The authors have declared no competing interest.

### Funding Statement

This project was funded by AIM-Ahead under project number 1OT2OD032581-02-303.

